# Are we ready for COVID-19’s Golden Passport? Insights from a Global Physician Survey

**DOI:** 10.1101/2020.11.25.20234195

**Authors:** P. Murali Doraiswamy, Mohan Chilukuri, Alexandra R. Linares, Katrina A. Bramstedt

## Abstract

**Introduction:** COVID-19 immunity passports could protect the right to free movement, but critics worry about insufficient evidence, privacy, fraud, and discrimination. We aimed to characterize the global physician community’s opinion regarding immunity passports.

**Methods:** Cross sectional, random stratified sample of physicians registered with Sermo, a global networking platform open to verified and licensed physicians. The survey aimed to sample 1,000 physicians divided among the USA, EU and rest of the world. The survey question on immunology asked physicians to offer their insights into whether we know enough about COVID-19 immunity and its duration to offer immunity passports at the present time.

**Results:** The survey was completed by 1004 physicians (67 specialties, 40 countries, 49% frontline specialties) with a mean (SD) age of 49.14 (12) years. Overall, 52% answered NO, 17% were UNCERTAIN, and 31% answered YES (*P* <0.05). EU physicians were more likely to sayYES but even among them it did not exceed 35% approval. US physicians (60%) were more likely to say NO.

**Conclusion:** Our findings suggest a current lack of support among physicians for immunity passports. It is hoped that ongoing research and vaccine trials will provide further clarity.

## INTRODUCTION

As of early January 2021, over 87 million people have been infected by SARS-CoV-2 worldwide[1] and COVID-19 immunity passports are being actively considered by several countries to facilitate travel and commerce while mitigating viral spread [2–10]. Immunity passports are envisioned as official travel documents which prove either vaccination or immunity for SARS-CoV-2. Immunity to a natural viral infection typically is a sequential, multi-dimensional process that comprises a non-specific innate response (to slow the progress of virus), antibodies that specifically bind to the virus and cellular immunity (T-cells to remove virus infected cells) [11]. A robust adaptive response may prevent re-infection by the same virus and may be detected by the presence of antibodies in blood [11].

Proponents argue immunity passports will protect the right to free movement and avoid penalizing those who are immune [2, 4–6, 9, 10]. Critics worry about insufficient evidence, privacy, passport fraud, and discrimination [3, 7–10]. There are also co-mingled issues of accuracy of antibody test results, virus mutation and re-infection, as well as vaccination [12–18]. For example, due to cross-reactivity, it is not yet known if antibody tests can accurately discriminate between past infections from SARS-CoV-2 versus infections from other common human coronaviruses (such as those that cause the common cold, Middle East Respiratory Syndrome or Severe Acute Respiratory Syndrome) [11]. Overall, COVID-19 presents a multiplicity of scientific and ethical complexity amid a global economy that is feeling the harsh effects of lockdowns and social distancing. Will immunity passports provide rescue? Despite the ongoing debate, relatively limited attention has been paid to the views of practicing physicians on the risks and benefits of immunity passports. The aim of this report is to present the opinions of global physicians.

## METHODS

### Study design and procedure

To characterize the opinions of physicians on this topic, we conducted a cross sectional, random, stratified sampling of verified physicians registered with Sermo, a secure digital platform for medical crowdsourcing and anonymous surveys.

### Study participants and sampling

The Sermo platform is exclusive to verified and licensed physicians and has over 800,000 registered physicians, of all specialties, worldwide. The survey sampled physicians over one week in September 2020 to get representation from the US, Europe and the rest of the world. As such this was an exploratory study and we aimed for a target sample size of 1000 doctors equally divided between USA, EU and rest of the world (RoW).

### Study instruments and measures

This was a cross-sectional, global, survey of doctors across 10 topics (such as digital surveillance, privacy, trust, risks) of which one question pertained to immunology and immunity passports. The instrument was refined after initial pilot testing in a sample of 25 doctors and then administered by an online survey. All questions asked doctors to give their opinions based on picking one answer from multiple choices. In addition, some quantitative and qualitative questions were also included. Results of other survey topics are being reported elsewhere. The survey question pertaining to immunity passports asked: “Digital immunity passports, based on antibody testing, are being considered to offer proof (e.g. via an app or QR code) that a person has developed lasting immunity to COVID-19 and hence can return to work or travel freely. In your opinion, do we know enough about COVID-19 immunity and it’s duration to offer such immunity passports at the present time?” Possible answers were YES, NO and UNCERTAIN.

### Ethical aspects

This anonymous survey was conducted following an online informed consent process. The survey results were de-linked to respondent’s personal identifiable information to create de-identified data. This analysis does not include any sensitive or identifiable data and was deemed exempt research by Duke University Medical Center’s institutional review board.

### Data analysis

Descriptive statistics were used to examine physicians’ characteristics and opinions by age group, gender, specialty and geographic region. Age grouping was done as younger versus older by age 50. We also examined differences by gender. Physicians directly diagnosing and treating COVID-19 were viewed as frontline (e.g. internal medicine, ICU, ED) whereas the rest were categorized for non-frontline (even though we recognize that all physicians may interact with or consult on COVID-19 patients). Chi-square and analyses of variance tests with p-values <0.05 were viewed as qualitatively different. Pairwise testing was done using Z-test or T-test, and chi-square were done without correction for small samples. Analysis was conducted using JMP Pro 15 from SAS as well as Protobi.

## RESULTS

### Socio-demographic characteristics

The survey was completed by 1004 physicians, across 67 specialties, from 40 countries. The mean (SD) age was 49.14 (12) years and approximately 49% were in frontline specialties (i.e., ICU, infectious disease, ED, hospitalists). 21% were female, 40% were male and 39% chose to not report their gender. 34% were from the US, 37% from EU and 29% from rest of the world.

### Physician opinions about immunity passports

#### Overall opinions

Among the entire sample, with regards to the utility of immunity passports, 52% of doctors answered NO, 31% answered YES, and 17% were UNCERTAIN (*P* < 0.0001).

#### Frontline status

When grouped by frontline status, there was no significant difference between frontline (30.0% YES, 53.6% NO, 16.3% UNCERTAIN) versus non-frontline (31.3% YES, 51.0% NO, 17.7% UNCERTAIN) physicians (*P* = 0.69).

#### Geographic practice region

When analyzed by major geographic region, physicians were still more likely to say NO than YES but there were regional differences (*P* = 0.01). US physicians (60.3%) were significantly more likely to say NO versus both EU (47.2%) (*P* < 0.001) and RoW physicians (49.5%) (*P* = 0.006) (Figure 1). A slightly higher percent of EU physicians (34.8%) said YES versus 25.9% of US physicians (*P* = 0.01) but in this regard EU did not differ from RoW physicians (31.1%).

**Figure 1.**
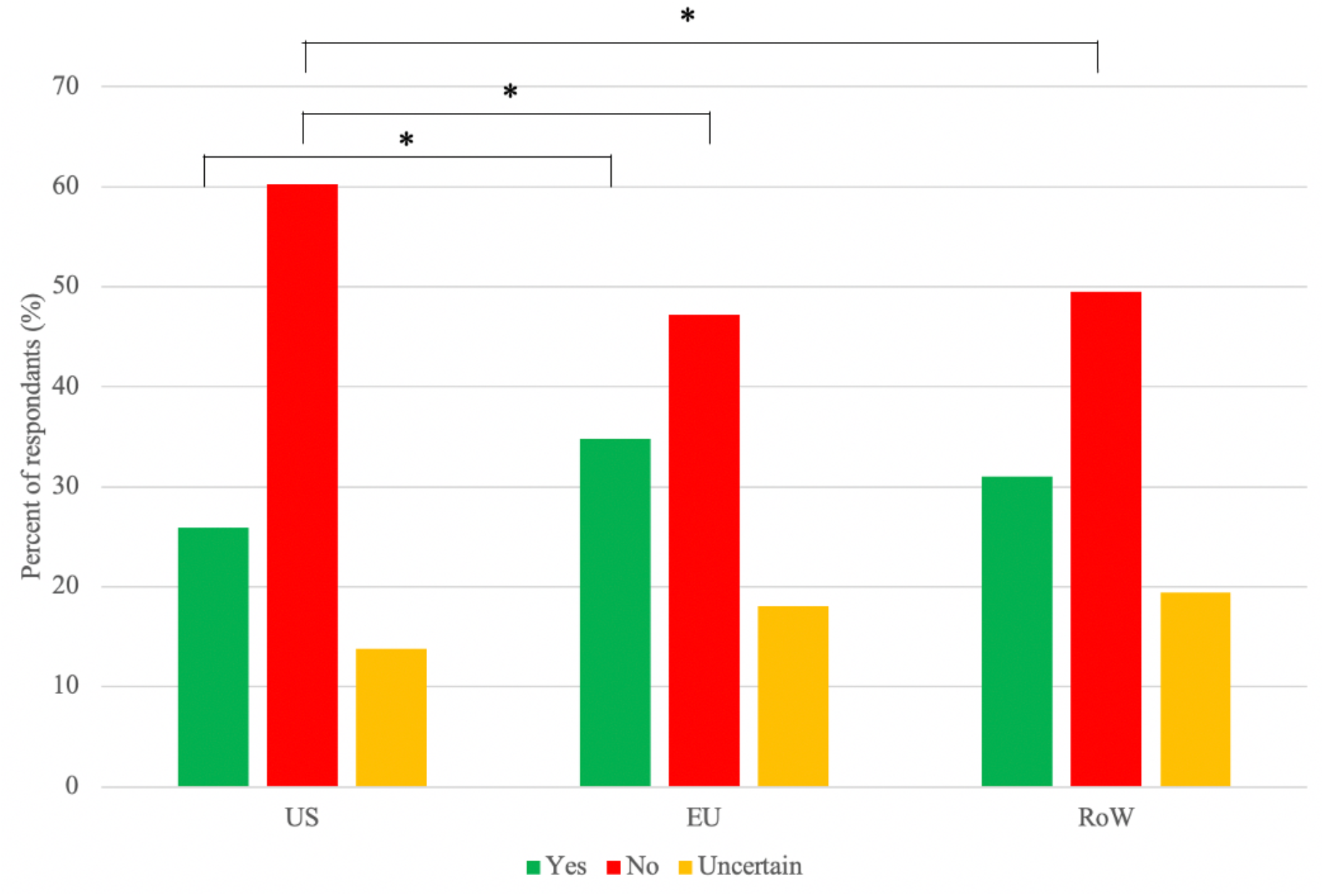
COVID-19 immunity passports: Physician responses by geographic region (*n* = 1004). ^*^ *P* < 0.05

#### Age effects

Older doctors also differed from younger doctors overall (*P* = 0.01). Older doctors (55.7%) were more likely to say NO than younger doctors (49.2%) (*P* = 0.04) and less likely to say YES (*P* = 0.003).

#### Gender effects

Among those who reported their gender, 65.2% of women said NO versus 56.5% of men (*P* = 0.037). Among EU female physicians, 67% said NO which was a higher number than for males respectively (*P* = 0.001). Among US female physicians, 70% said NO and this number rose to 80% among US older female physicians. While this number was higher than the respective percentages for males, it was not statistically different. The female versus male difference was not seen among the RoW physicians.

## DISCUSSION

Immunity passports are being actively considered by several countries and the subject of considerable debate [2–10]. In that context, our survey provides the first global insight into how practicing physicians think about this issue. Our findings suggest a current lack of support among the global physician community for immunity passports. Even in the EU and Asia, where physicians and researchers have been working with COVID-19 for a longer duration than other parts of the world, physicians in these regions generally believe there is not enough evidence about COVID-19 antibodies to warrant immunity passports.

We speculate there could be many reasons why physicians are skeptical. The validity of this possibility is supported by recent conflicting findings about the accuracy of antibody tests, the duration of COVID-19 immunity, and how variations in test terminology can impact user behavior [3, 7, 10, 16–18]. These data are also consistent with cautionary comments from the WHO that antibody tests (including rapid immunodiagnostics) to SARS-CoV-2 need further research validation at both test accuracy and effectiveness levels. Suboptimal test accuracy could result in false positives or false negatives, both of which could result in harm. Further, people with antibodies may wrongly assume they are risk free and ignore public health advice [11]. Another explanation for our findings is that the current burden of COVID-19 cases, including the second wave hitting much of the world, has focused physicians more on managing the severe, daily problems of morbidity and mortality [19]. From an ethics perspective, vaccine development and movement of people across borders are both community goods, but for a physician workforce faced with daily demands of patients with COVID-19, treatment is a real-time priority, and the idea of patient travel is more focused on sending them home from the hospital healthy (rather than sending them off for travel). Sporadic, global reports of viral mutation and patient re-infection [12–14] could contribute to our results, as both mutation and re-infection widen the scope of need for immunity investigations, as well as vaccine development. Reflecting on influenza, there are numerous strains, but the yearly vaccines are of limited scope. If this pattern emerges with COVID-19, there could be cohorts of those with ‘immunity passports’ but would these be ‘golden’ (valuable) if there is a strain mismatch [3]?

### Study strengths and limitations

To our knowledge, this is the first global survey to investigate the opinions of physicians about immunity passports. The survey benefited from a relatively large sample drawn from diverse specialties and practice settings across 40 countries. The use of Sermo, a global platform of verified and licensed physicians, allowed us to recruit verified frontline and nonfrontline practicing doctors. There are also some limitations with our survey. These include the relative absence of respondents from developing regions (e.g. Africa), sampling bias (e.g. people registered on a platform), response biases (e.g. degree to which they are interested in topic may have influenced their participation) and confounding effects of variables not measured. Also a cross-sectional survey cannot determine causality or predictive validity – indeed the risks and benefits of immunity passports may not be fully known for decades. Thus, while our findings must be viewed as preliminary, they may provide valuable insights for policy makers.

Overall, a healthy global economy requires a healthy population (those cured of COVID-19 and those prevented from acquiring it). This population will need to move across regions for both leisure and commerce. Even though physicians are not yet ready for immunity passports, the International Air Transport Association is moving forward with the idea [4] and plans pilot testing of their Travel Pass in late 2020. It is hoped that the ongoing vaccine trials, analyses of serology data from millions of patients worldwide, and genomic and molecular studies of spike protein variations will provide answers to many of the unsolved COVID-19 immunology questions [5, 15].

## CONCLUSION

Our survey provides foundational insights into how global physicians think about COVID-19 immunity passports. These findings call for further research to establish the accuracy and effectiveness of antibody testing. As a growing proportion of society gets vaccinated, there will be renewed calls for ‘immunity passports’ based on vaccination status. Further research is warranted to elucidate the duration of immunity conferred by the various COVID-19 vaccines, whether vaccinated individuals could still infect others, and how best to ensure that any future division of society by immunity status does not worsen existing societal inequities.

## Data Availability

Data will be made available upon approval for publication in a peer reviewed journal.

## Acknowledgments

The authors thank Sermo and the physicians who took part in the survey.

## Author Contributions

PMD drafted the paper, and ARL conducted analysis. PMD and ARL verified the underlying data. All authors aided in interpretation of results and manuscript edits.

